# Utility of gene tumor expression of *VEGF, FOXM1*3* and *CD-133* on diagnosis and prognosis of brain gliomas

**DOI:** 10.1101/2020.08.15.20175166

**Authors:** Iris Angélica Feria-Romero, Bárbara Nettel-Rueda, Marco Antonio Rodríguez-Florido, Ignacio Félix-Espinoza, Luis Castellanos-Pallares, Jesús Cienfuegos-Meza, Sandra Orozco-Suárez, Jesús A. Chavez, Consuelo Escamilla-Nuñez, Gerardo Guinto, Horacio Márquez-González, Carlos Rodea-Ávila, Israel Grijalva

## Abstract

**Objective:** This paper seeks to quantify the normalized expression of transcripts *FOXM1*3, VEGF, CD133*, and *MGMT* and their relation with the histopathological and molecular diagnosis and with the probability of estimating tumor progression-free survival of gliomas.

**Methods:** A cohort of patients was made up of patients aged over 18 years with a histological and molecular diagnosis of gliomas from the year 2011 to 2018. The patients had a complete tumor resection. Patients with high-grade glioma received adjuvant management (temozolamide and radiotherapy). Clinical and imaging follow-up was carried out periodically to identify the time of progression free survival (PFS).

**Results:** Ninety-one patients (age range, 18–85 years) comprised the study cohort with a predominance of males. The expression of *FOXM1*3, VEGF*, and *CD133* allowed the differentiation of astrocytomas grade II from GBM. ROC curves proved statistically significant in the GBM model (*p* < 0.05), demonstrating greatest sensitivity with *FOXM1*3* (91%), and greatest specificity with *VEGF* (93%). The age-adjusted Cox multivariate model established that a PFS_50%_ of 25 months corresponds to a median value of 5.3 for *VEGF* and 0.42 for CD133.

**Conclusions:** The normalized expression of transcripts FOXM1*\*3, VEGF*, and *CD133* allow us to estimate the probability of PFS, especially in gliomas grades II and IV; likewise, their overexpression defines the diagnosis of GBM.

**Authorship:**

1. Substantial contributions to conception and design (IAFR, BNR, MARF, GG, IG), acquisition of data (IAFR, BNR, MARF, IFE, LCP, JCM, SOS, JAC, CRA), analysis and interpretation of data (IAFR, BNR, MARF, JCM, SOS, CEN, HMG, IG).
2. Drafting the article (IAFR, BNR, MARF, IFE, LCP, SOS, JAC, IG), revising it critically for important intellectual content (IAFR, JCM, CEN, GG, HMG, CRA, IG)
3. Final approval of the version to be published (IAFR, BNR, MARF, IFE, LCP, JCM, SOS, JAC, CEN, GG, HMG, CRA, IG).

## INTRODUCTION

Glioma comprise a group of primary tumors that are characterized by the presence of abnormal glial-lineage cells, but only the diffuse astrocytic glioma develops the most aggressive variety of this group of lesions: Glioblastoma (GBM).^1^

The updated 2016 edition of the World Health Organization (WHO) Classification of Tumours of the Central Nervous System (CNS) (2016 WHO CNS), in addition to the histopathological study, includes: the evaluation of the protein Ki67; the mutation of the gene isocitrate dehydrogenase 1 (IDH-1, G395A) and of its protein (R132H); methylation of the gene promoter methyl-guanine-O6-DNA methyltransferase (MGMT), and the codeletion of 1p19q, which allows the identification of oligodendrogliomas (O) together with the nuclear expression of the transcriptional regulator ATRX.^2 – 5^

Despite the fact that the correct diagnosis of gliomas permits the adequate selection of the standard treatment, it is known that in some cases there is a discrepancy between the behavior predicted by the histological classification and the therapeutic response of the local tumor and survival.^6^

The biological processes of mitosis, angiogenesis, cellular immaturity, and even resistance to chemotherapy presented by gliomas can be evaluated with molecular markers that allow for determining the tumor’s malignancy as well as the patient’s prognosis.^7^

Currently, there are markers that have been used in the diagnosis and prognosis of different types of cancer, as well as methylation of the promoter of the gene MGMT, which has been studied as a predictive and prognostic factor of response to treatment with adjuvant and concomitant radiotherapy and chemotherapy in high-grade brain tumors^8 – 10^, as well as the forkhead box M1 transcription factor (FOXM1), the vascular endothelial growth factor (VEGF), and CD133, whose predictive usefulness has apparently not been explored sufficiently, although they also have been studied in gliomas^11 – 14^.

Following standard treatment of patients with a diagnosis of brain gliomas according to the update 2016 WHO CNS, the objective of this paper was to identify whether the expression of FOXM1 isoform 3 (FOXM1*3), VEGF, CD133, and MGMT would allow for estimating the probability of progression free survival (PFS), as well as learning whether the expression of the four cited transcripts could be related to the histopathological and molecular diagnosis according to the 2016 WHO CNS.

## METHODS

### Patients and obtaining the sample

The project was approved by the National Committee for Scientific Research of the Mexican Institute of Social Security (IMSS) and was conducted from 2011 through 2018. A cohort of patients with a diagnostic suspicion of brain gliomas were integrated into the study protocol.

The patients were recruited in the Neurosurgery Department, of Specialty Hospital “Bernardo Sepúlveda”, IMSS Century XXI National Medical Center, in Mexico City, once they had accepted to participate voluntarily in the protocol and signed the letter of informed consent. Patients of both sexes were included, over the age of 18, and with a diagnosis of low- and high-grade gliomas by histology and molecular tests of diagnostic certainty, and were recruited from 2011 through 2018. All cohort patients had undergone a complete tumor resection, and those who presented high-grade data also received adjuvant management according to the Stupp protocol.^15^

Time zero was considered as the day that the surgical procedure was performed. Follow-up was clinical and through imaging. The initial imaging study of the imaging follow-up was carried out 24 h after surgery, and subsequently every 6 months; clinical follow-up was performed monthly after the patient left the hospital during the first 3 months and afterward every trimester or semester, depending on the diagnosis of low grade or high grade, until the clinical or imaging suspicion of tumor progression was identified. The outcome was based on the period of PFS, defined as the time in months from the moment of tumor resection of the detection of tumor activity, through the detection of secondary tumor activity, and until the deterioration of neurological conditions confirmed through imaging studies. The clinical and demographic characteristics of the patients were obtained from the clinical history or by means of a direct interview with the patient.

The histopathological diagnosis was carried out separately by three neuropathologists and was considered definitive when at least two of them agreed with subsequent molecular test results. The grade of malignancy was determined mainly by the expression of astrocytomas, oligodendrogliomas, and oligoastrocytomas, and was further determined by the loss or presence of the nuclear expression of ATRX and the presence or loss of the regions that identify the arms of chromosomes 1p and/or 19q. Ependymomas (E) grade II were confirmed by the expression of the protein epithelial membrane antigen (EMA).

During the surgical procedure, one to three fragments were obtained from different representative regions of the tumor, and were immediately frozen with liquid nitrogen and later stored at –70°C until time of processing. The brain tissue of reference was obtained from the healthy cortex of both brain hemispheres, through the donation of autopsies of five individuals (*n* = 10) who died of different causes, without evidence of any neurological disease. Genomic DNA and RNA were extracted from frozen samples.

### Quantification of transcripts *IPO8, FOXM1*3, VEGF, CD133*, and *MGMT* by qPCR

Gene transcripts *IPO8, FOXM1* isoform 3 (*FOXM1*3*), *VEGFA, MGMT*, and *CD133* were amplified from complementary DNA and detected with the TaqMan reagent and real time thermocycler of LightCycler 2.0 (Roche, Germany), using proprietary designs. The values of Cp were interpolated in the concentration curve corresponding to each gene. Lastly, the linear values were normalized with the Housekeeping gene (HG) *IP08*. The regions selected for detection were the following: *IPO8*, NM_00119 0995.1, region 453–599; *FOXM1*3*, NM_202003.2, region 1,160–1,305; *MGMT*, NM_002412.4, region 319–450; *VEGFA*, NM_001025366.2; region 1,208–1,377, and *CD133*, NM_006017.2, region 1,814–1,967.

### Identification of 1p/19q by qPCR

The genes *LRRC47* (1p36.32), *SOAT1* (1q25.2), *MAN2B1* (19p13.13), and *SYNGR4* (19q13.33) of the samples and controls (lymphocytes from healthy volunteers) were amplified from genomic DNA and detected with the TaqMan reagent and real time thermocycler of LightCycler 2.0 (Roche, Germany), using proprietary designs. The relative expression of the problem genes (*LRRC47* and *SYNGR4*) as compared to their reference genes (*SOAT1* and *MAN2B1*) were obtained with the Pffaffi equation.^16^ The regions selected for detection were the following: *LRRC47*, NC_000001.11, region 3795769–3795915; *SOAT1*, NC_000001.11, region 179335786–179335641; *MAN2B1*, NC_000019.10, region 12655782–12655904, and *SYNGR4*, NC_000019.10, region 48373525–48373385.

### Identification of *IDH1* mut

Genomic DNA of 17 tumor samples was amplified the 6,412–6,806 region of the gene *IDH1* (NG_023319.2), afterward, amplicons were sequenced to identify the heterozygotes of the mutation G6756A. Lastly, the same genomic DNA region was amplified, and detected with the SYBER Green reagent and real time thermocycler of LightCycler 2.0 (Roche, Germany) to obtain the denaturalization Temperature (Tm) of the samples. The wild-type variant (wt, G6756G) was identified when Tm ≥79.80 and the mutated variant (mut, G6756A) was identified when Tm < 79.80.

### Identification of Ki67 and ATRX by immunohistochemistry

Immunohistochemistry detection was performed using five micron-thick sections, antibodies to Ki-67 (Biocare, USA) and ATRX (Abcam, USA), and staining with DAB system (Biocare, USA).

### Statistical analysis

A descriptive analysis was carried out to determine the parametric distribution of the data, represented as median interquartile range. Analysis of the median differences of the transcripts in the different tumor types was conducted by means of the Kruskal Wallis and Mann Whitney U tests, where the value of p ≤0.05 was considered a significant difference. The linear discriminant model identified that the expression of transcripts *FOXM1*3, VEGF, CD133*, and *MGMT* normalized by HG I*PO8* and the combinations among them achieved the differentiation of the diagnosis of glioma according to WHO CNS 2016. The optimal cut-off point of the transcripts was identified using the proposed probability of a GBM, employing a multiple logistic regression model. Later, a bootstrap analysis was proposed to identify the optimal cut-off point, sensitivity, specificity, and area under the curve (AUC). To evaluate the PFS and the risks associated with the transcripts, a univariate analysis and the Cox multivariate model was proposed. Lastly, survival curves were constructed to determine the probability of being free of tumor progression.

## RESULTS Clinical Data

Ninety-one patients comprised the cohort-of-study. The cases analyzed ranged in age between 18 and 85 years, the median age being 54 years. The median age of the patients upon stratifying the grade of malignancy was progressively higher. The patient with a diagnosis of pilocytic astrocytoma (PA I) was 33 years old; cases with glioma grades II, III, and IV were a median 34 years of age [range, 29.25–46.25 years], 40 years [range, 34.5–43 years], and 62 years [range, 50–70 years], respectively. Male patients were predominant, especially in the group of grade II gliomas. One lobe was affected in 58.2% of cases, mainly in the frontal lobe, presenting a history of gliomas grade II (45.7%). The most frequent cellular lineage was astrocytic (80/91 cases, 87.9%) and the main histopathological molecular diagnosis was GBM (57/91 cases, 62.6%) (table 1).

**Table 1.**
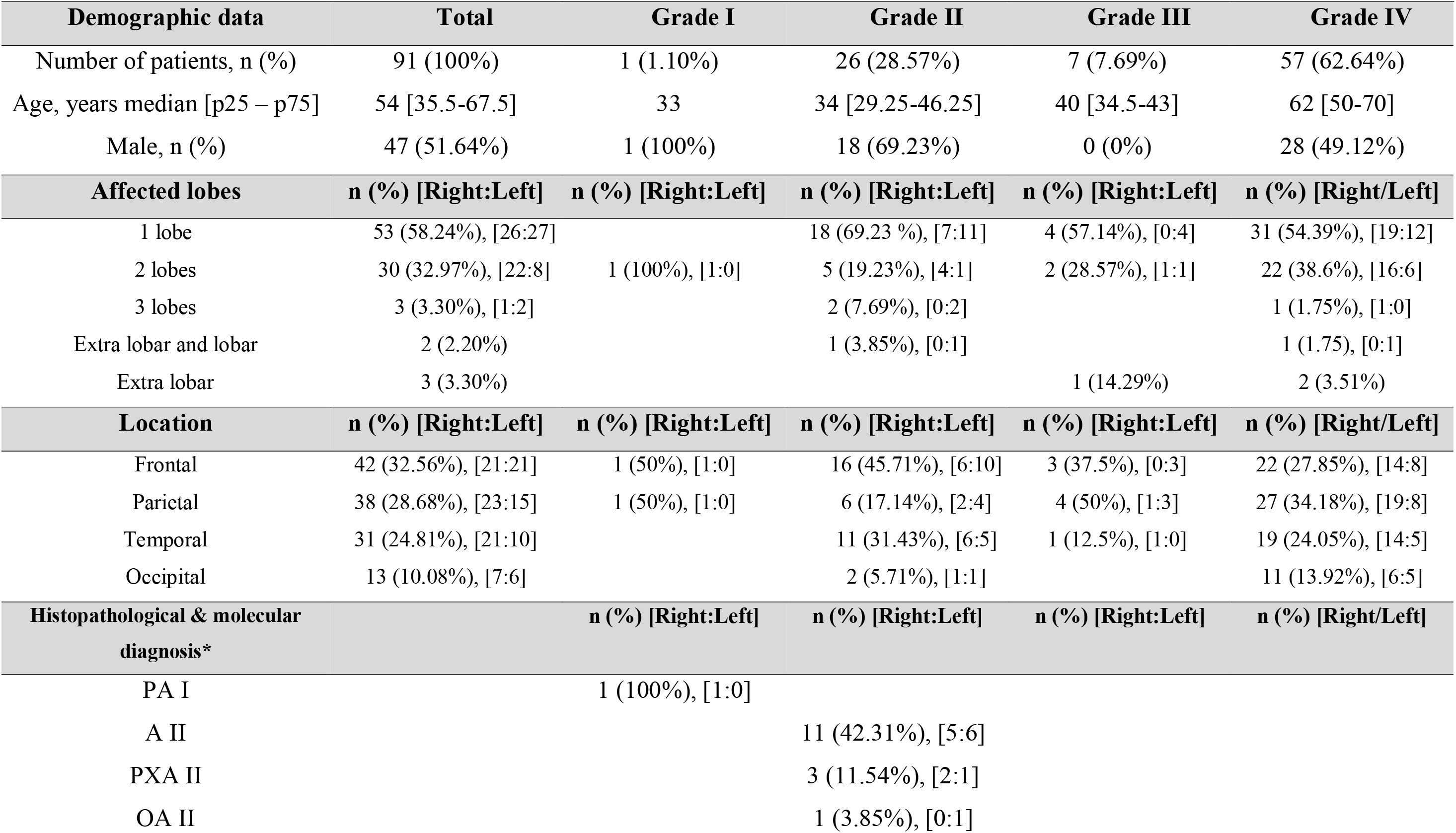
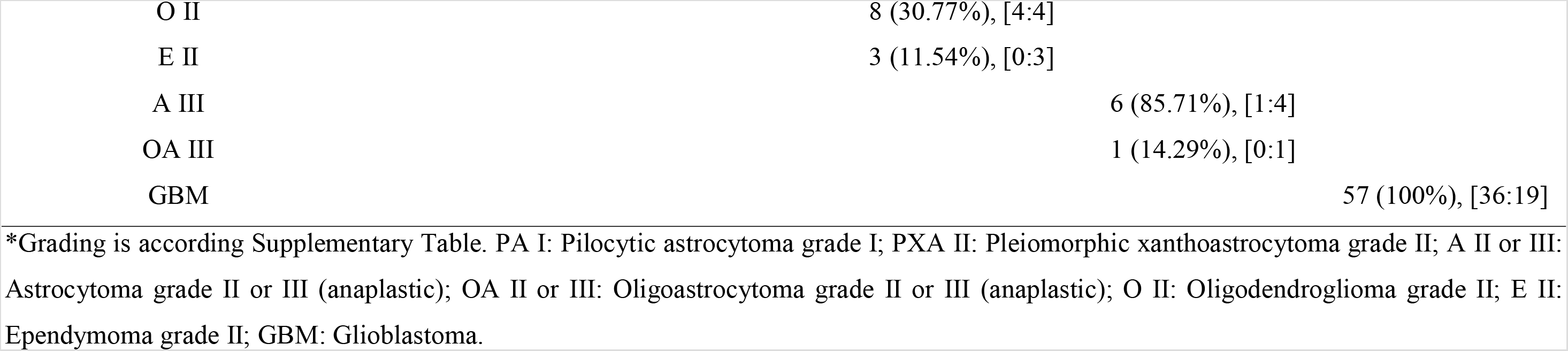
Demographic data, location, laterality and histopathological and molecular diagnosis of patients with gliomas

The mutated form of *IDH-1* predominated in grade II and III astrocytomas, as well as in grade II ependymomas. Regarding GBM, the sub-classification was determined by the native or mutated form of *IDH-1*, in which 37/57 *IDH1* wt GBM (65%) were identified. The histopathological and molecular characteristics of the patients’ tumors are shown in table 2.

**Table 2.**
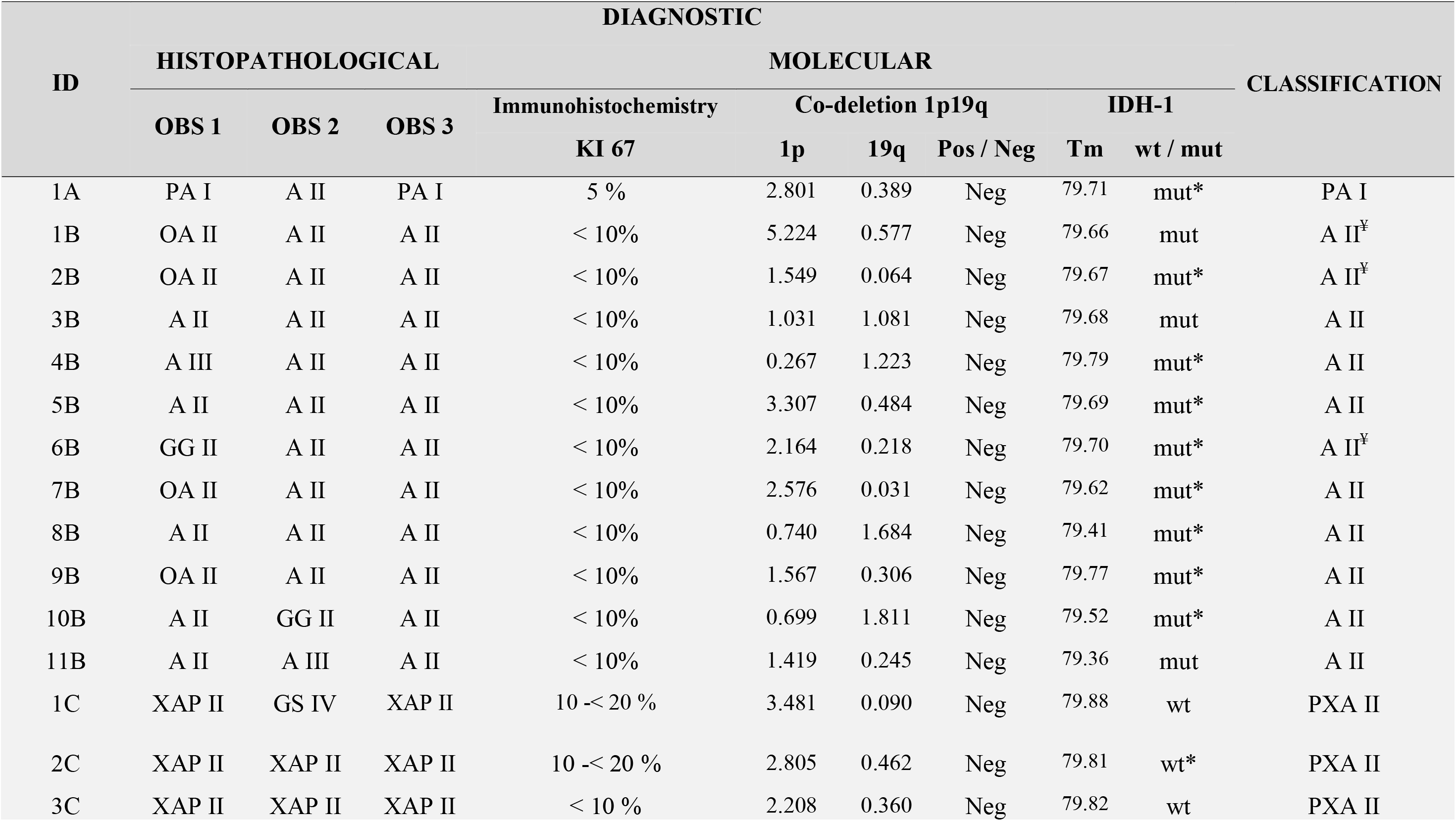
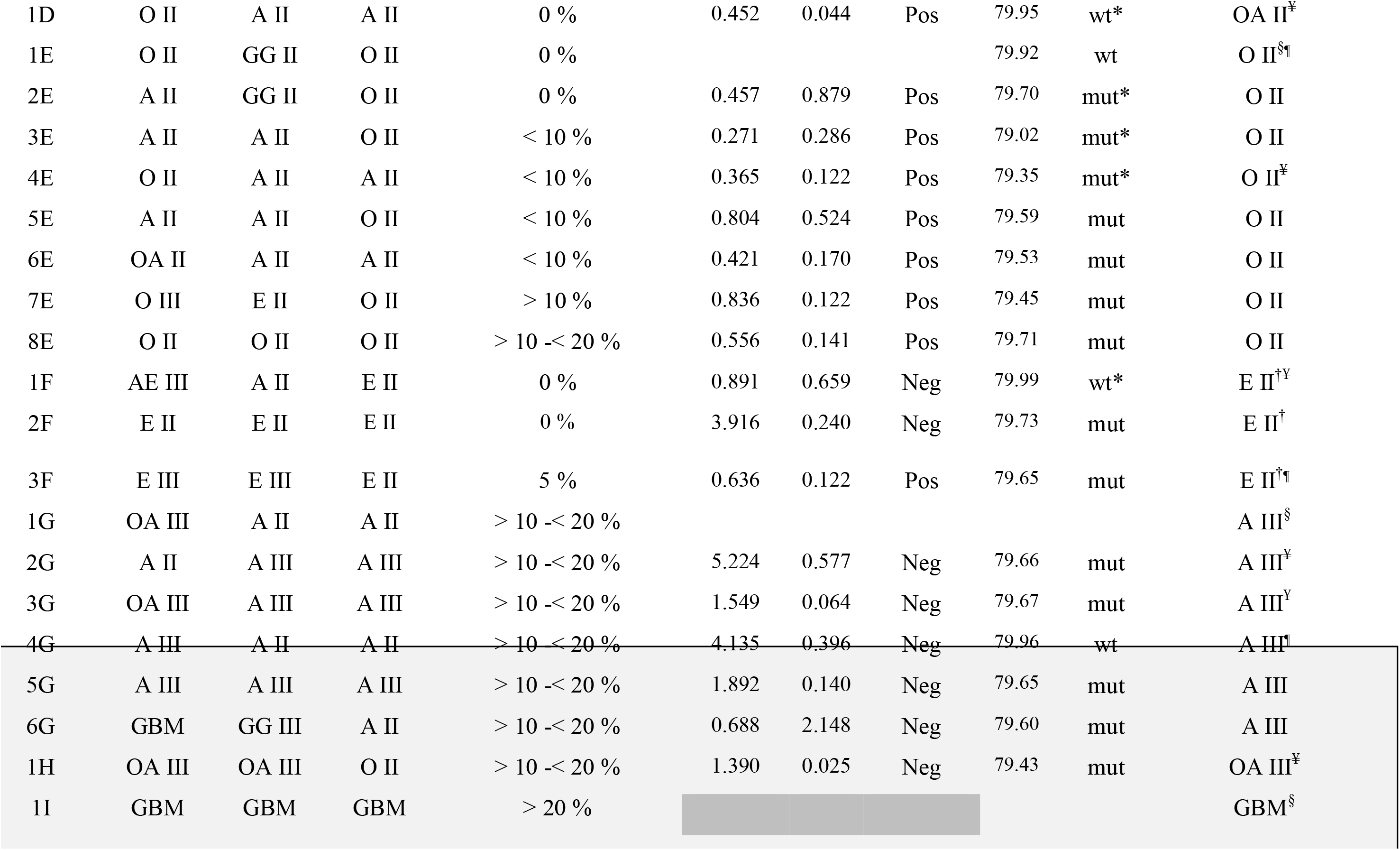
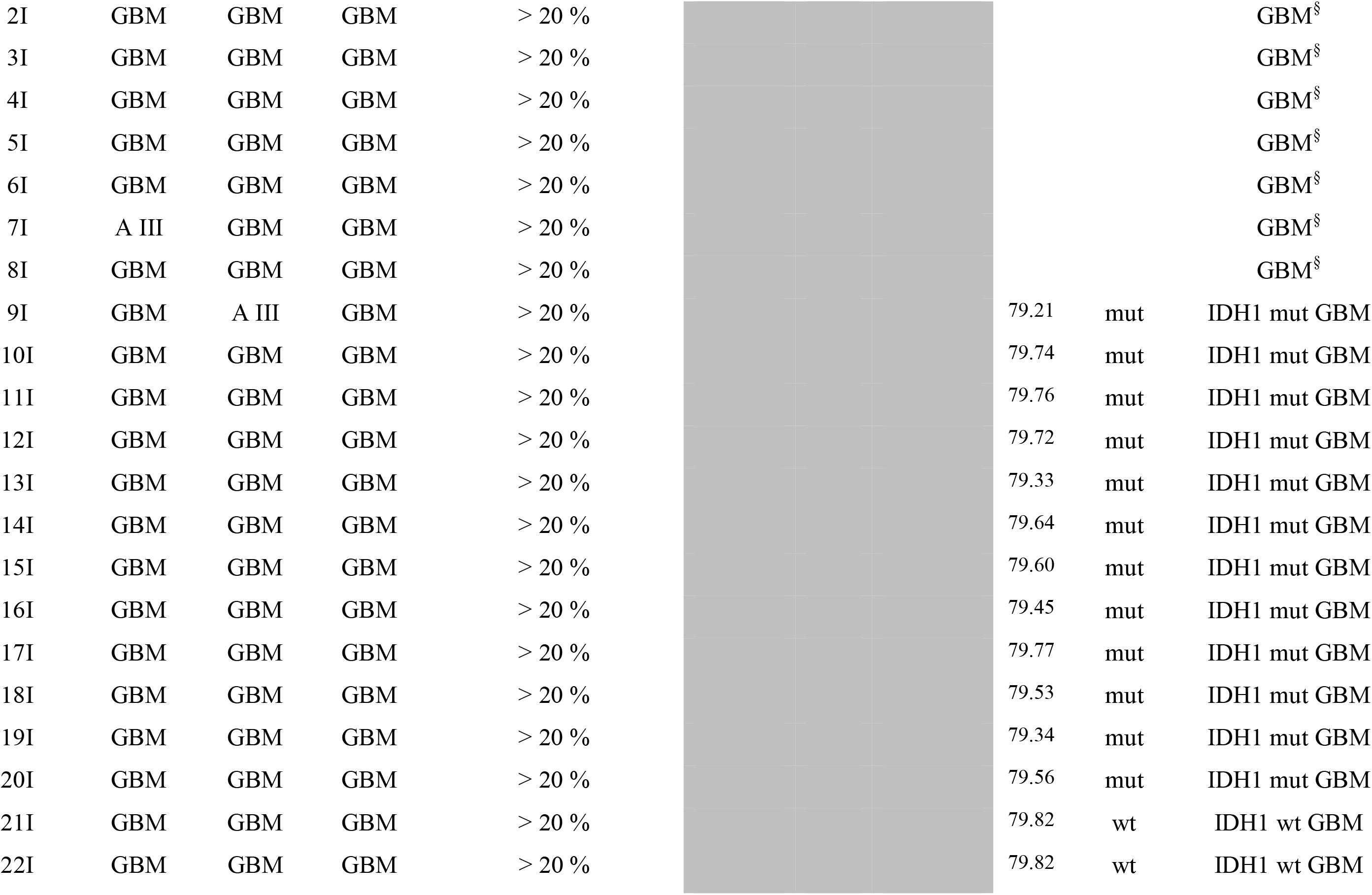
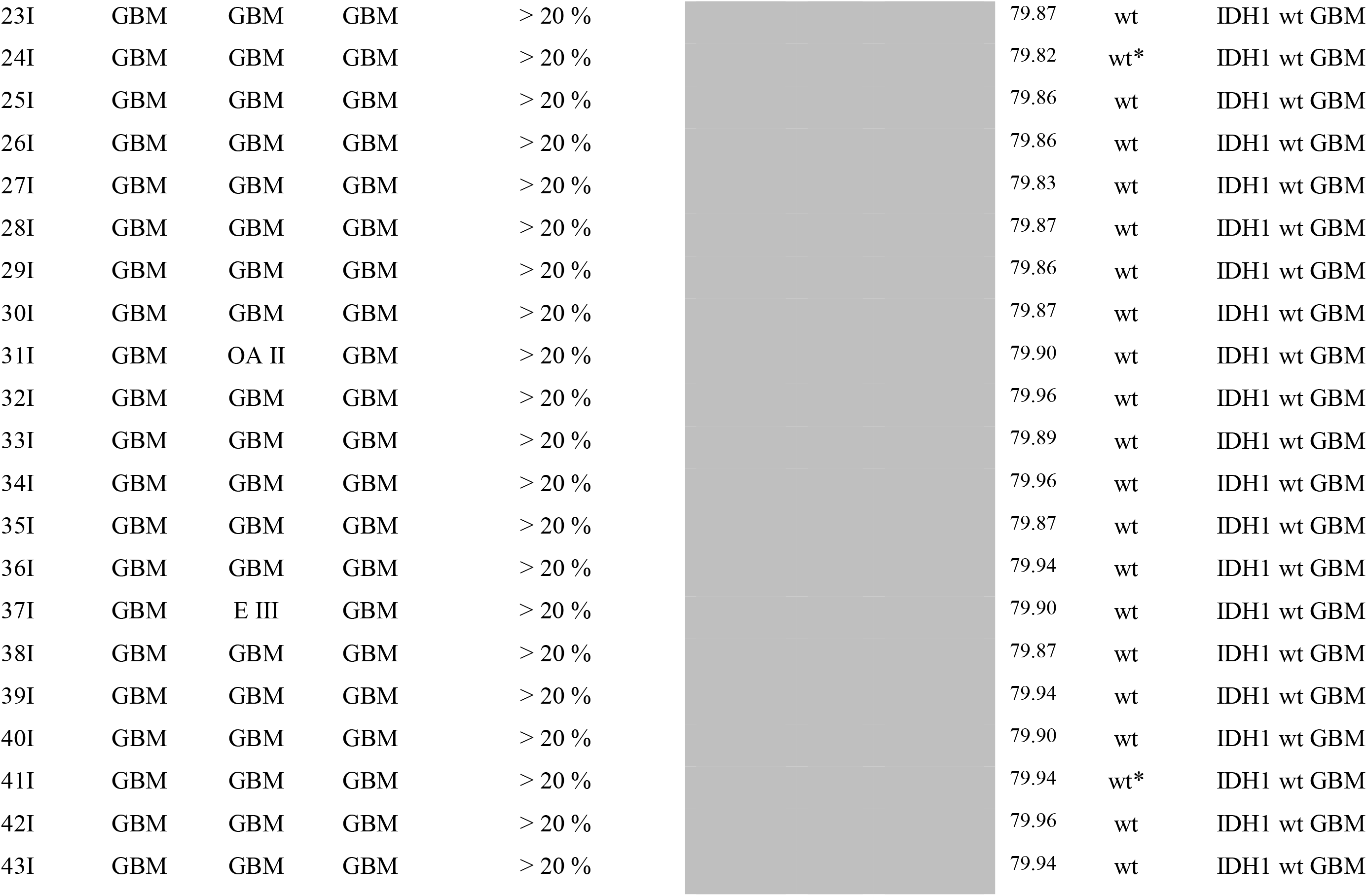
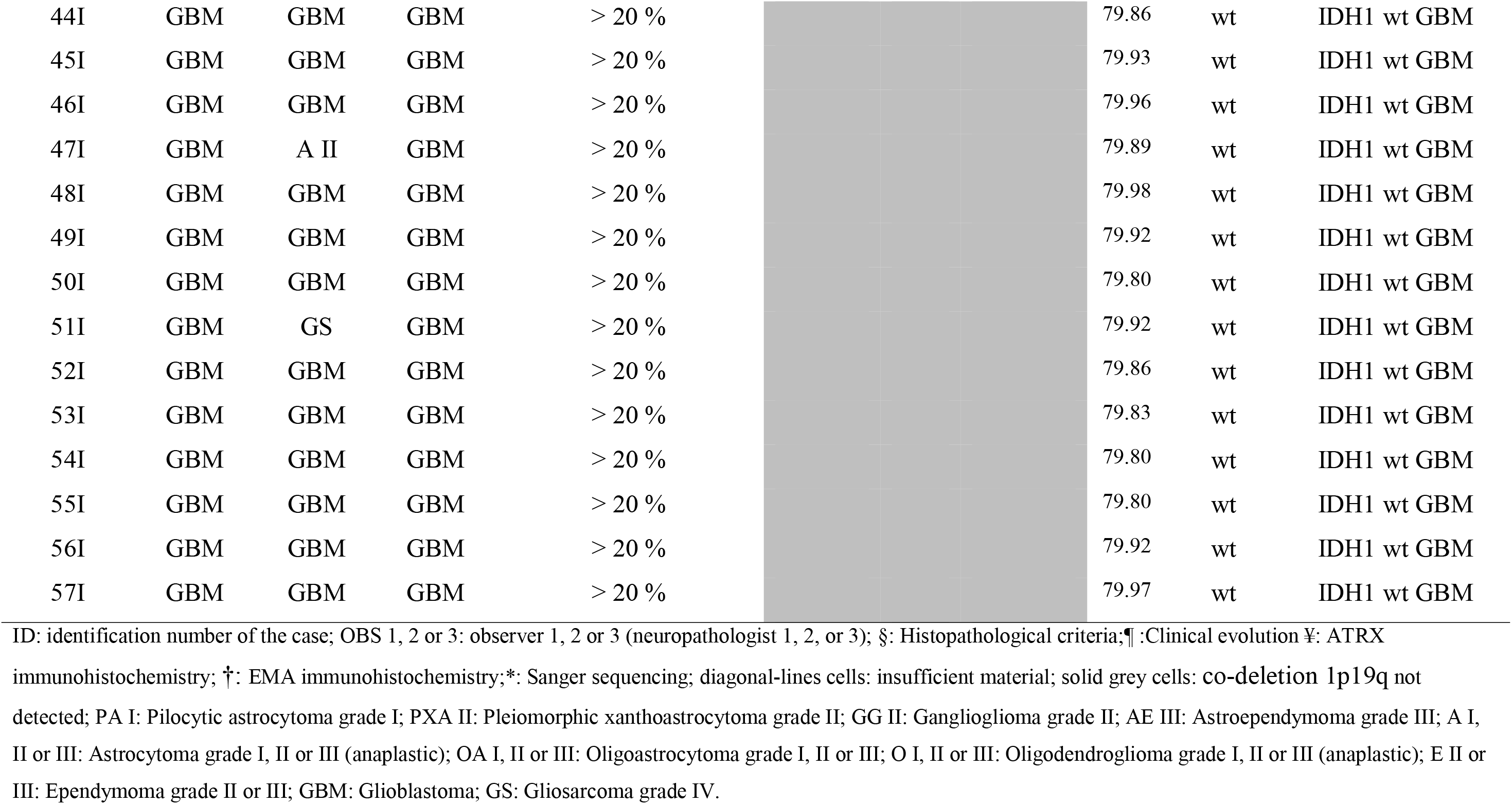
Molecular and histopathological criteria in classification of gliomas

### Normalized Expression of the Transcripts

Comparison of medians among all of the grouped gliomas showed differences in the expression of *FOXM1*3* (*p* < 0.0001), *VEGF* (*p* < 0.0001), and *CD133* (*p* < 0.0001), but not with transcript *MGMT* (*p* = 0.5952). Later, employing the corresponding post-hoc test, significant differences were observed in the expression of *FOXM1*3* between the control group (Cx) and groups A II, O II, A III and the two GBM groups; and between the GBM *IDH1* wt and groups A II, pleomorphic xantoastrocytoma (PXA) II, OII, E II, and A III (figure 1A). In *VEGF*, statistical differences were observed between the two GBM groups and the Cx groups, that is, A II, PXA II, and O II (figure 1B). Regarding *CD133*, significant differences were found between the GBM *IDH1* wt group and Cx, A II, PXA II, O II, and A III groups (figure 1C). Upon studying the *MGMT* transcript, no significant differences were observed among the groups (figure 1D). None of the four transcripts could differentiate primary and secondary GBM. On comparing the median and the p25-p75 expression range of the groups, it was observed that *FOXM1*3* allowed for differentiating between the Cx group and grade II gliomas (A II, PXA II, O II, and E II), as well as the A II group and GBM *IDH1* wt. Regarding *VEGF*, GBM *IDH1* wt, and Cx groups and grade II gliomas were differentiated; also, groups A II and A III were separated. With *CD133*, the two GBM groups and all of the remaining study groups were differentiated, with the exception of E II (table 3).

**Figure 1.**
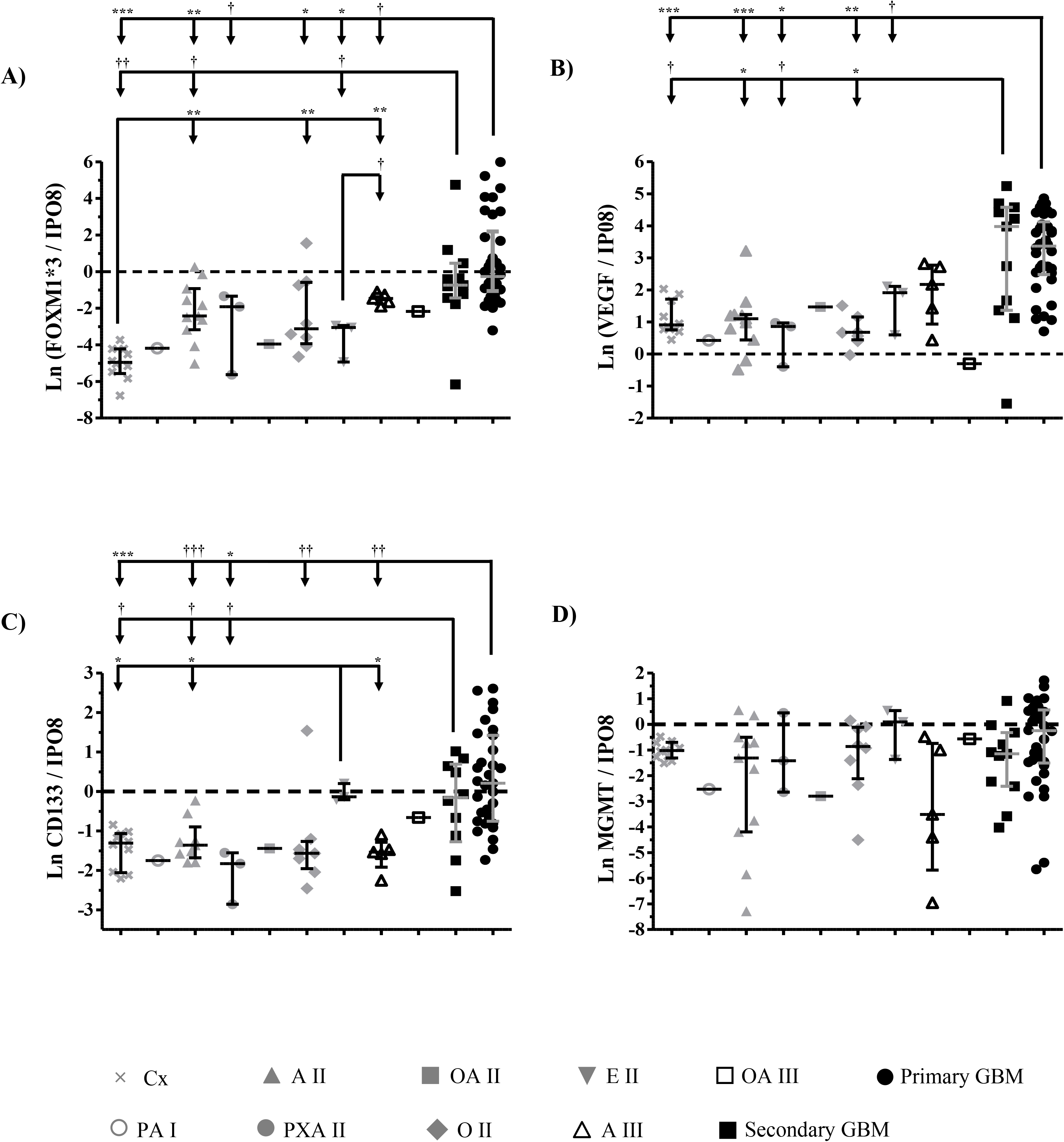
Expression of transcripts *FOXM1*3, VEGF, CD133*, and *MGMT* in healthy cortex and the gliomas of the cohort. A) *FOXM1*3*, B) *VEGF*, C) *CD133*, and D) *MGMT*. The values were normalized with *IPO8*; these were transformed to their Ln and were analyzed with the Mann Whitney *U* test. \**p* <0.01, † *p* <0.05, ** *p* <0.001, †† *p* <0.005, *** *p* <0.0001, and ††† *p* <0.0005. Cx, control; A II, astrocytoma grade II; OA II, oligoastrocytoma grade II; E II, ependymoma grade II; OA III, oligoastrocytoma grade III; GBM, glioblastoma; PA I, pilocytic astrocytoma grade I; PXA II, pleomorphic xantoastrocytoma grade II; O II, oligodendroglioma grade II; A III, astrocytoma grade III.

**Table 3.** Expression of normalized transcripts in different groups of gliomas

| Group | n | FOXM1*3 | VEGF | CD133 | MGMT |
| --- | --- | --- | --- | --- | --- |
|  |  | Median [p25-p75] |  |  |  |
| Cx | 10 | **0.01 [0.00 – 0.01]** | **2.48 [2.16 – 4.74]** | **0.27 [0.15 – 0.33]** | 0.36 [0.28 – 0.46] |
| A II | 11 | **0.09 [0.06 – 0.30]** | 3.01 [1.87 – 3.39] | 0.22 [0.19 – 0.28] | 0.48 [0.18 – 0.61] |
| PXA II | 3 | 0.15 [0.08 – 0.20] | 2.36 [1.52 – 2.50] | 0.16 [0.11 – 0.19] | 0.24 [0.16 – 0.91] |
| O II | 8 | 0.05 [0.03 – 0.63] | 1.97 [1.70 – 3.18] | 0.21 [0.17 – 0.25] | 0.42 [0.21 – 0.86] |
| E II | 3 | 0.05 [0.03 – 0.50] | 6.77 [4.29 – 7.51] | 0.87 [0.84 – 1.05] | 1.10 [0.68 – 1.41] |
| A III | 5 | 0.20 [0.15 – 0.28] | 8.78 [4.19 – 15.21] | 0.21 [0.10 – 0.21] | 0.03 [0.03 – 0.43] |
| *IDH1* mut GBM | 11 | 0.48 [0.27 – 1.14] | 53.42 [4.63 – 90.40] | 0.79 [0.41 – 1.76] | 0.32 [0.10 – 0.59] |
| *IDH1* wt GBM | 34 | **0.76 [0.37 – 6.29]** | **29 [12.59 – 56.18]** | **1.08 [0.45 – 2.67]** | 0.78 [0.23 – 1.71] |

Bold range: Values with same line cell but not overlap in different malignancy grade. Cx: control group; A II: Astrocytoma grade II; PXA II: Pleiomorphic xanthoastrocytoma grade II; O II: Oligodendroglioma grade II; E II: Ependymoma grade II; A III: Astrocytoma grade III (anaplastic); GBM: Glioblastoma.

### Usefulness of the Expression of the Transcripts in the Correct Classification of the Gliomas

The application of the discriminant model with the values transformed into the natural logarithm of the normalized expression of transcripts *FOXM1*3, VEGF, CD133*, and *MGMT* to identify agreement with the histopathological diagnoses was able to differentiate GBM as a sole group. In 40/45 of cases (88.9%) diagnosed histopathologically molecularly, such as GBM, the same diagnosis was maintained, with a 6.8% probability of error. One hundred percent of the healthy brain cortexes conserved the same diagnosis with a probability of 53.5% of remaining in the same group. Last, 7/13 of the cases (53.8%) diagnosed as A II maintained their initial diagnosis, with a probability of less than 55.61% of remaining in that category. In the rest of the groups, expression of the transcripts was not in agreement with their initial diagnosis.

### Identification of the Optimal Cut-off Point

The multiple logistic regression analysis identified that 95% of patients with an initial histopathological molecular diagnosis of GBM were classified correctly from the predicted probabilities (*p* < 0.5 and *p* ≥0.5), with a sensitivity of 92.7%, a specificity of 97.4%, and an area under the ROC curve of 96.6%.

Later, from the ROC curves, the expression was analyzed for each transcript separately. The normalized transcripts of FOXM1*3, *VEGF*, and *CD133* were statistically significant in the GBM model (*p* < 0.05). Greatest sensitivity was obtained with *FOXM1*3* (91%), followed by *CD133* (85%) and *VEGF* (76%). Greatest specificity was obtained with *VEGF* (93%), followed by *CD133* (80%) and *FOXM1*3* (76%). In the case of *MGMT*, despite presenting a specificity of 80%, sensitivity was 47% (figure 2).

**Figure 2.**
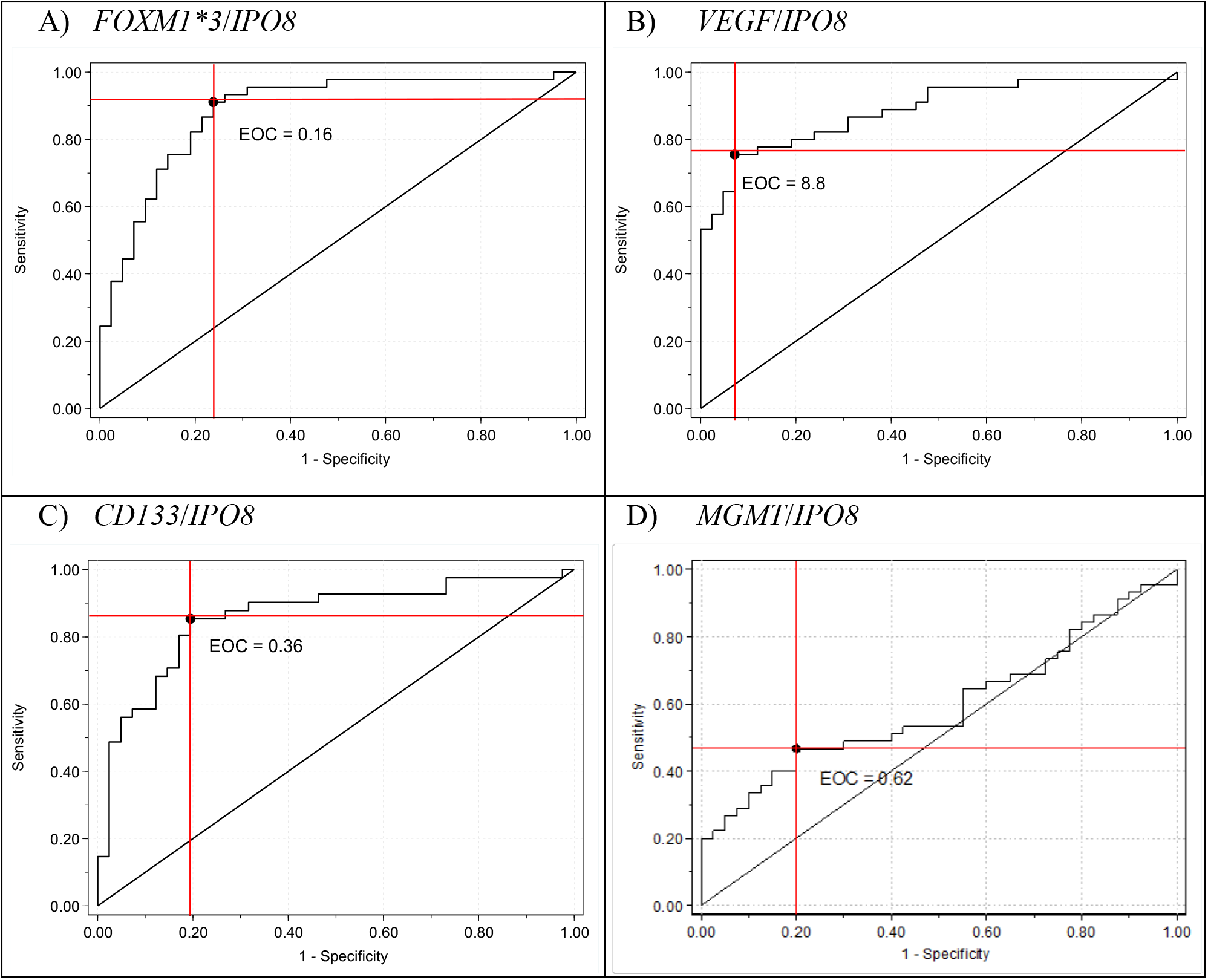
Estimation of the cut-off point for the diagnosis of GBM. A) *VEGF*: EOC = 6.5, Sensitivity = 0.90, Specificity = 0.92, and AUC ROC curve = 0.94; B) *FOXM1*3*: EOC = 0.21, Sensitivity = 0.79, Specificity = 0.72, and AUC ROC curve = 0.58. A logistic regression model and bootstrap analysis were used. EOC: Empirical Optimal Cut-off point; AUC ROC curve = 0.81; C) *CD133*: EOC = 0.33, Sensitivity = 0.69, Specificity = 0.76, and AUC ROC curve = 0.66, and D) *MGMT*: EOC = 0.09, Sensitivity = 0.86, Specificity = 30.6, and AUC ROC curve: Area Under the Curve on a Operating Characteristic curve.

### Progression free survival of glioma

Analysis of tumor progression-free survival (PFS) was carried out with the follow-up and outcome information of 51/93 patients in the cohort (54.8%) with different histopathological diagnoses. In the univariate statistical analysis, a significant HR was observed for tumor regrowth with the normalized value of transcripts *FOXM1*3, VEGF*, and *CD133*, as well as the clinical variable of age. In the multivariate analysis, only transcripts *VEGF* and *CD133* conserved a significant HR with respect to tumor regrowth (table 4).

**Table 4.** Univariate and multivariate analysis related with progression free survival of glioma and FOXM1*3, VEGF and CD133 transcripts expression

| Variable | Univariate* |  | Multivariate* |  |
| --- | --- | --- | --- | --- |
|  | HR (95%CI) | P value | HR (95%CI) | P value |
| *FOXM1\*3/IPO8* | 3.3 (2.9, 3.8) | 0.001 |  |  |
| *VEGF/IPO8* | 4.2 (3.2, 5.9) | 0.001 | 3.9 (2.9, 5.7) | 0.010 |
| *CD133/IPO8* | 4.3 (3.1, 6.5) | 0.003 | 3.9 (2.9, 6.0) | 0.021 |
| *MGMT/IPO8* | 3.5 (2.7, 4.9) | 0.057 |  |  |
| Age (years) | 2.8 (2.7, 2.8) | 0.009 |  |  |

\* 51 patients included in the Cox multivariate model of proportional hazards regression; HR, hazard ratio.

On the other hand, it was observed that 50% of the patients who presented the median and the range of p25-p75 of the normalized value of age-adjusted *VEGF* of 5.3 [2.6–31.2], had the corresponding PFS_50%_ of 25 months [16.9–36]. With respect to *CD133*, the value of the median and the p25-p75 range was 0.42 [0.22–1.23] and the corresponding PFS_50%_ was 25 months [20.1–30] (figure 3).

**Figure 3.**
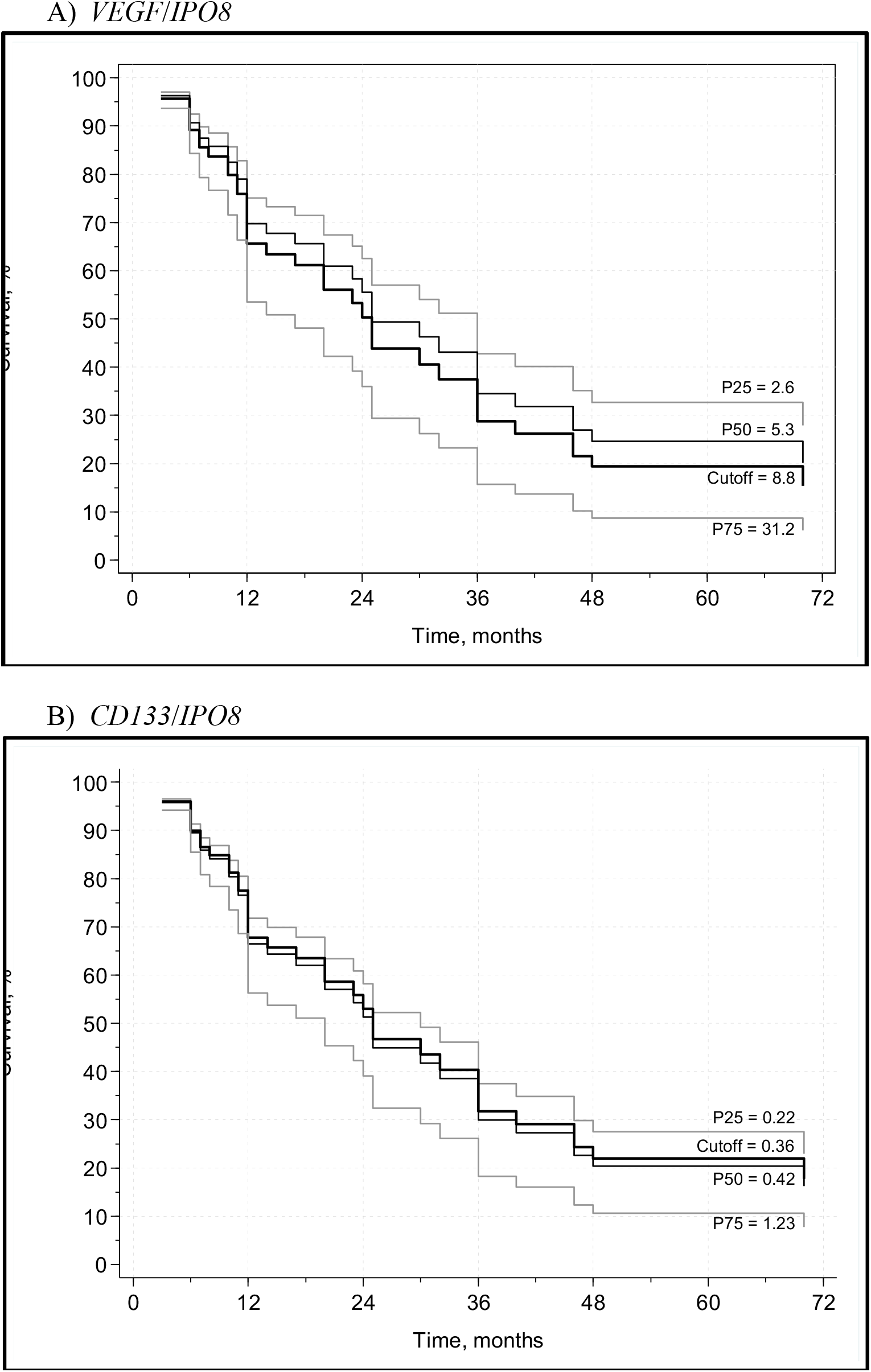
Tumor progression-free survival of patients with a diagnosis of GBM. The analysis was carried out with the Cox multivariate model of proportional hazards regression and the expression of transcripts with significant risk *VEGF* and *CD133*, normalized with *IPO8* and adjusted for age.

## DISCUSSION

The present work demonstrated how the quantification of transcripts *FOXM1*3, VEGF*, and *CD-133* can contribute to determine the definitive diagnosis of the gliomas, as well as to identify tumor progression-free survival probabilistically, especially in GBM.

In this study’s cohort, males were predominant, as was frontal lobe localization; the main lineage was astrocytic, and the primary diagnosis was GBM. This is in agreement with the results of the last report published by the Central Brain Tumor Registry Of The United States (CBTRUS)^17^.

### Histopathological and Molecular Diagnosis

Since 2016, the proposed WHO classification of tumours of the CNS update has also included the identification of Ki67, ATRX or EMA^1^, the mutation of the gene *IDH-1* and the 1p19q co-deletion.^5^. As opposed to what is usually done identify the mutation of IDH-1 (R132H) by immunohistochemistry^18^ or the co-deletion of 1p19q by means of the FISH technique^19^ in this work, both genetic alterations were identified by the qPCR technique. Hence, the result was contingent on a significant number of cells presenting the alteration or not.

On the other hand, selection of the transcripts (*FOXM1, VEGF, CD133*, and *MGMT*) was based on prior studies that identified changes in the expression of these transcripts in the gliomas, such as the most representative of the clinical, histopathological and biological characteristics of the GBM (cellular proliferation, angiogenesis, gliogenic stem cell population, and resistance to TMZ) in addition to the metabolic pathways involved.^11 20 – 22^ Lastly, transcript *IPO8* was selected as an endogenous gene, since it does not present gliogenic stem cell population important changes of expression in gliomas with different grades of malignancy compared to the normal brain.^23^

*FOXM1*3* was selected, because the expression of this isoform in healthy cortex is practically absent, which is more desirable for the identification of low-grade gliomas. Our results are similar to those reported by van den Boom J et al. (2003)^24^, who conducted a comparative analysis of the mRNA of astrocytomas of differing grades of malignancy, employing the qPCR technique. In this work, the authors observed significant differences between A II vs. A III and A II vs. GBM, but not between A III vs. GBM and primary GBM vs. secondary GBM; however, the authors did not make this distinction in the isoform.

In the case of the transcript *VEGF*, Plate KH et al. (1992)^21^, employing the Northern blot technique, there was a significant increase observed (50 times) of RNAm *VEGF* in GBM compared with A II; in addition, van den Boom J et al. (2003)^24^, using the qPCR technique, observed significant differences between A II vs. GBM and A III vs. GBM, but not between A II vs. A III and primary GBM vs. secondary GBM. In contrast to the reports previously cited, in this work the expression of mRNA *VEGF* normalized with *IPO8* permitted differentiating between low-grade (A II) and high-grade astrocytomas (A III and GBM) which agrees with Yang et al., (2016)25 and other authors with different techniques of VEGF evaluation.^26 – 28^

Transcript *CD133* has been identified in high-grade astrocytomas (A III and GBM), and their principal relation is to Glioma-forming Stem Cells (GSC), as well as the presence of neurospheres in the primary tissue of patients with GBM.^20^ In addition to what was previously reported, in this work we identified the range of expression of CD133 in healthy cerebral cortex, as well as its significant increase in GBM.

Finally, in contrast with the three transcripts cited previously, the expression of the MGMT transcript does not permit identifying the lineage nor the grade of malignancy of the gliomas studied.

### Prognosis of the Patient in Relation to Their Tumor Progression-Free Survival

When we applied the discriminant model with the study transcripts to the patients’ samples, we observed that the three groups that best met the statistical criteria necessary to maintain their original classification were Cx, A II, and GBM. However, only the healthy cortex group was clearly differentiated from the remaining groups. In group A II, there were two cases that could be reclassified as Cx with a PFS of ≥ 36 months, and two cases that could be reclassified as GBM presented a PFS at < 36 months. In the GBM group, one case that could be reclassified as A III at the cut-off of this work has not presented tumor activity (24 months). In the remaining cases, a probable reclassification was not consistent with the clinical evolution of the patient. Worth noting is a previous work that applies the discriminant model and the normalized expression of transcripts *COL1*A2, *POSTN, NNMT*, and *DCN*) selected by prior analysis of the microarrangements of 46 GBM and one gliosarcoma, whose main objective was to find the discriminant score (ColSBE) that would identify GBM of good (sub-type GLE) and poor prognosis (sub-type GHE).^29^

Without taking into account the expression of the MGMT transcript given that it was not deemed meaningful for prognosis, pursuant to that reported by Tang K et al. (2012)^30^ who showed that MGMT protein expression level was not a prognostic factor, as opposed to other publications that do attest to this^31 32^, in the present work, a congruent and consistent prognosis was established, mainly in three groups: 1) Patients with grade II gliomas of oligodendrocytic lineage (O II and OA II) with low expression of *FOXM1*3*, that is, between 0.00 and 0.04, *VEGF* <2.6, and *CD133* between 0.00 and 0.01 independently of the result of *IDH1* who presented a PFS of >70 months; 2) Patients with grade II gliomas (astrocytoma, oligodendroglioma, oligoastrocytoma, or pleomorphic xantoastrocytoma) with expression of *FOXM1*3* between 0.05 and 0.1, *VEGF* ≤2.6, *CD133* between 0.02 and 0.22 independently of the state of *IDH1*, which presented a PFS of ≥30 months. In this group, the exception was observed in the AP I in which the *IDH1* mut was detected and that evolved into a gliosarcoma at 12 months. 3) Patients with a GBM diagnosis that showing high expression of VEGF (>30) and CD133 (>1.2), in 80% of the cases a PFS of <4 months was observed. Our results agree with several papers considering lower levels of these biomarkers on longer PFS^25 26 28^ and higher levels on shorter PFS.^25 26 28 33 – 35^

While our results suggest a tight relation between the levels of expression of the tumor markers *VEGF, FOXM1*3, CD133*, and the PFS, this does not mean that they will lack the known classic markers (methylation of the promotor of *MGMT, IDH1* mut, and co-deletion 1p-19q). These new markers are added to the diagnostic armamentarium to define the prognosis, particularly in dubious cases.

There are important limitations to our study. Due to the strict selection criteria, a small number of patients was included; a larger sample size could have improved the results obtained. Another important limitation was the number of patients lost in follow-up. In conclusion, the increased expression of transcripts *FOXM1, VEGF*, and *CD133* did permit estimating the probability of tumor progression-free survival, especially in gliomas grade II and IV; on the other hand, an increase in the expression of these transcripts can define GBM independently from the 2016 WHO CNS diagnosis.

## Data Availability

All relevant data are within the manuscript and its supporting information file.

## Acknowledgments

We would like to thank Dr. Rocío Lorena Arreola Rosales from the Department of Pathologic Anatomy for her invaluable help by providing the histopathological material. We also would like to thank to David Ramos Ponce for his important technical assistance.

## Notes

Competing interests: Authors have no competing interests to disclose.

### Competing Interest Statement

The authors have declared no competing interest.

### Funding Statement

The present study was funded by Fondo de Investigacion en Salud del Instituto Mexicano del Seguro Social grant (FIS/IMSS/PRIO/10/012).

### Author Declarations

The project was approved by the National Committee for Scientific Research of the Mexican Institute of Social Security (IMSS), registration number 2010-785-042.

